# Temporal Recalibration in Schizophrenia: A Compensatory Timing Trap?

**DOI:** 10.1101/2025.08.13.25333560

**Authors:** Ali Aytemur, Ayşen Esen Danacı, Osman İyilikci

**Author notes:** Corresponding author: Ali Aytemur, Manisa Celal Bayar University, Faculty of Humanities and Social Sciences, Department of Psychology, Manisa / Turkey.

## Abstract

**Background:** Schizophrenia is characterised by widespread neural dysconnectivity and impaired temporal coordination. Despite these pervasive disruptions, patients can maintain coherent perception and functional behaviour, particularly during remission. This paradox is underexplored in the literature, which has primarily focused on symptom emergence. We propose a dual-role hypothesis of sensorimotor temporal recalibration, a process known for adapting to temporal discrepancies in sensorimotor integration. We hypothesise that temporal recalibration may serve both as a compensatory mechanism mitigating neural incoherence and as a trigger for positive symptoms.

**Method:** To investigate its compensatory role, we compared 20 clinically stable schizophrenia patients to 20 matched healthy controls using a visuomotor temporal order judgement task following adaptation to varying sensorimotor asynchronies (0, 150 and 300 ms).

**Results:** Our findings revealed that patients with schizophrenia exhibited significantly greater temporal recalibration compared to controls. Across participants, recalibration was stronger for longer delays. No group differences were observed in just noticeable differences (JNDs) suggesting comparable task difficulty and temporal precision.

**Conclusions:** These findings suggest that patients not only have an intact temporal recalibration mechanism but also may engage it more to counteract neural delays and preserve temporal coherence. We further propose that overreliance on this adaptive mechanism may paradoxically contribute to symptom emergence under conditions of temporal instability by distorting the perceived order of actions and their sensory consequences. This dual-role hypothesis offers a novel perspective for understanding how the same temporal mechanism can sustain perceptual coherence, yet under certain conditions, contribute to the breakdown of causality and agency, which underlie control delusions and hallucinations.

## Introduction

Schizophrenia involves impaired communication^1,2^ and disrupted temporal coordination^3,4^ between large-scale brain networks. These disruptions are thought to contribute to the fragmented perception and disrupted sense of agency, the experience of control over voluntary actions and their sensory consequences, in schizophrenia. However, current theories mainly focus on the mechanisms underlying symptom manifestation and often overlook how patients maintain relatively preserved perception and agency when neural timing remains disrupted. Therefore, revealing the mechanisms compensating for temporal coordination abnormalities is essential for understanding perceptual functionality in schizophrenia. In addition, exploring compensatory mechanisms is crucial for identifying factors that may interrupt these processes and contribute to the positive symptoms. In this study, we propose a novel hypothesis that extends current dysconnectivity models by addressing two key issues: how relatively coherent perception is maintained despite temporal incoherence, and how this very compensatory process may contribute to symptom onset under certain conditions.

Temporal coordination is crucial for integrating information across different systems into unified perceptual experience. Studies in healthy participants showed that asynchronous inputs can be integrated into a unified perception by adaptively shifting their timing to overcome temporal discrepancies, a process known as temporal recalibration.^5–7^ Especially, sensorimotor temporal recalibration proposed to be a key mechanism for supporting functional interaction with the environment by compensating temporal discrepancies between motor commands and their sensory consequences.^6^ In schizophrenia, numerous studies have reported greater levels of structural and functional connectivity abnormalities^1^ that contribute to temporal asynchrony across neural systems.^4^ Notably, these disruptions persist even in clinical remission and shared with patients’ unaffected first-degree relatives or with high-risk individuals.^8–13^ In addition, schizophrenia has been associated with demyelination related efference copy delays^14–16^ and slowed or less efficient visual information processing^17–21^ which can further contribute to sensorimotor timing asynchronies that threaten the integrity of perception unless actively corrected. These patterns of abnormalities may specifically require increased levels of temporal adjustments for perceptual coherence in the visuo-motor domain since both action initiation and visual processing rely on the integrity of these affected networks. Therefore, investigating whether schizophrenia patients rely on heightened recalibration to maintain perceptual coherence could provide novel insights into the adaptive mechanisms that counterbalance neural dysconnectivity and asynchrony.

While temporal recalibration has been considered as an adaptive process, its potential overuse in schizophrenia may have disruptive consequences. Sensorimotor temporal recalibration studies in healthy participants showed that perceived temporal order of actions and their sensory consequences can be reversed when the adapted temporal asynchrony between them is reduced or removed, leading participants to perceive sensory consequences as occurring prior to their initiating actions. ^6,22,23^ Therefore, transient reductions in the adapted temporal asynchronies between motor and sensory components can diminish perceived causality between them and lead to attributing one’s agency to external sources^24^ as sensory events preceding their initiating actions cannot plausibly be self-generated. In schizophrenia, transient reductions in adapted sensorimotor asynchronies may arise from acute changes in neural processing underpinned by heightened arousal, dopaminergic dysregulation or transient neural synchrony, that have been linked to psychosis.^4,25–28^ Therefore, this can mimic the findings in healthy individuals and excessive reliance on temporal recalibration mechanism may increase vulnerability to positive symptoms under conditions of transient neural processing change by increasing the risk of temporal order distortions.

We propose a dual role for temporal recalibration in schizophrenia. That is, while overreliance on temporal recalibration mechanism may generally support perceptual stability by compensating increased neural asynchrony, it may also paradoxically increase the risk of temporal order disruptions and potentially contribute to positive symptoms, particularly under conditions of increased neural gain or information speed. This hypothesis bridges dysconnectivity models of schizophrenia with mechanistic accounts of perceptual timing and agency. In the current study, we focused on testing the compensatory aspect of this hypothesis. Specifically, we investigated whether clinically stable schizophrenia patients show heightened sensorimotor temporal recalibration compared to healthy controls, as a potential mechanism for mitigating timing discrepancies between motor actions and their sensory consequences.

## Methods

### Participants

The study included 20 individuals diagnosed with schizophrenia and 20 healthy control participants. Participants with schizophrenia (8 females,12 males; age: M = 38.9, SD = 9.8) were recruited from the outpatient unit of the Manisa Celal Bayar University Hospital Psychiatry Clinic. Diagnosis was established using the Structured Clinical Interview for DSM-IV Axis I Disorders (SCID-I),^29^ and all patients were receiving outpatient treatment at the time of participation. Inclusion criteria for the schizophrenia group were: (1) no change in psychiatric medication within the past month, (2) no inpatient psychiatric treatment within the past three months, (3) no history of electroconvulsive therapy, (4) no diagnosis of neurological disorders, and (5) no alcohol or substance addiction. Symptom severity was assessed using the Scale for the Assessment of Positive Symptoms (SAPS)^30,31^ and the Scale for the Assessment of Negative Symptoms (SANS).^32,33^ The control group (8 females, 12 males; age: M = 39.6, SD = 9.3) was recruited from the local community and matched to the patient group on age, gender, and years of education. Inclusion criteria for controls included: (1) no history of psychiatric or neurological disorders, (2) no family history of schizophrenia, schizoaffective disorder, or bipolar disorder, and (3) no history of alcohol or substance addiction. Four participants from the schizophrenia group were initially excluded due to chance-level performance at the largest SOA (375 ms) in the baseline condition. Their responses across other SOAs were similarly unmodulated, resulting in near-flat psychometric functions indicative of non-systematic or inattentive responding. These were replaced by newly recruited individuals who met the same inclusion criteria. One of the replacements was also excluded and replaced because of the same reason above, resulting in a final sample of 20 patients with schizophrenia. Demographic characteristics and clinical data for both groups are summarized in Supplementary Table 1.

The authors assert that all procedures contributing to this work comply with the ethical standards of the relevant national and institutional committees on human experimentation and with the Helsinki Declaration of 1975, as revised in 2013. All procedures involving human subjects/patients were approved by Clinical Research Ethics Committee of the Faculty of Medicine at Manisa Celal Bayar University (Approval No: 429). All participants provided written informed consent prior to participation.

### Methods and Materials

For the experimental task, participants sat in front of a computer screen (144Hz, 27 inch). The temporal recalibration task was presented using OpenSesame 4.0^34^ on a high-performance gaming computer (MSI MEG Trident X, 12th Gen Intel Core i7, NVIDIA RTX 3080, 32 GB RAM) running Windows 11. Because the timing was critical for this study, participants made button presses using a custom-built grip handle silent button connected to an Arduino micro-controller (< 2 ms latency).^35^ Silent button was used for preventing button press noise to be perceived as the auditory outcome of their actions.

### Temporal recalibration task

Temporal recalibration task involved adaptation and test phase.^6,22,23^ Participants were instructed to make 5 consecutive button presses with approximately 1 second interval in each trial. The first 4 button presses constituted the adaptation period, and each button press led to the presentation of a white square with 0 ms (baseline condition), 150 ms (short delay condition) or 300 ms (long delay condition) delay depending on the condition. The fifth button press served as the testing period. On this final press, participants were presented randomly with the visual stimulus (white square) at one of the 7 stimulus onset asynchronies (SOAs): −112 (on average, physically before the button press), 0, 75, 150, 225, 300 and 375 ms. On the testing period participants performed a temporal order judgement task where they were asked whether white square appeared before or after their button press. Each SOA was presented for 18 trials in each condition resulting in 126 trials in total for a condition. To present the visual stimulus before the button presses on the testing period, we kept a running average of the participants button press intervals from the adaptation period and predicted their 5^th^ button press timing. We subtracted 75 or 150 ms (first two SOAs in the stimulus set) from the predicted timing of the 5^th^ button press. Using this approach, we were able to present on average 112 ms (SD = 42 ms) before the participants’ button press in “before” trials. This average was calculated across all conditions and groups to generate a fixed SOA for ‘before’ trials and to avoid introducing bias when fitting psychometric functions. Delay conditions (0, 150 and 300 ms delay duration conditions) were counterbalanced as no delay condition or delay condition first^24^ and short and long delay conditions were also counterbalanced within themselves as short delay (150 ms) or long delay (300 ms) condition first. This procedure resulted in these four scenarios (No Delay, Short Delay, Long Delay), (No Delay, Long Delay, Short Delay), (Long Delay, Short Delay, No Delay), (Short Delay, Long Delay, No Delay). Participants completed a practice session involving 6 trials of 375 ms, 0 ms and visual stimulus before button press SoAs in the No-delay condition before the main task started.

## Results

### Point of subjective simultaneity

Each participants’ proportions of “after” responses were calculated for each SoA and experimental condition and modelled by fitting psychometric functions in the form of a cumulative Gaussian distribution function using Python 3.11 and the Psignifit module.^36^ Participants’ point of subjective simultaneities as the 50% cut-point of all trials (PSS: the temporal difference needed for button press and visual stimulus to be perceived simultaneous) and just noticeable differences (JND: representing the interval where 27% and 73 % after responses were given)^23^ were calculated for each condition. PSS shifts in the direction of the inserted delay (e.g.: PSS shift from No-Delay Condition to Delay Conditions) has been considered as temporal recalibration.^6,22,23^ Hence, temporal recalibration effects for Short Delay and Long Delay Conditions were calculated by subtracting PSS value in the No Delay Condition (baseline condition) from the PSS values in these delay conditions.

### Temporal recalibration analysis

To examine group differences in temporal recalibration, we analysed PSS shifts from the No-Delay (Baseline) condition to the two delay conditions (Short and Long Delay conditions), which served as a direct index of temporal recalibration effect. A mixed-design ANOVA was conducted on temporal recalibration effects with group (Schizophrenia Patients and Healthy Controls) as a between subject variable and condition (Short and Long Delay) as a within subject variable. This analysis revealed a significant main effect of group [*F* (1,38) = 5.187, *p* = 0.028, *ηp*^2^ = 0.120] indicating that schizophrenia patients showed significantly higher levels of temporal recalibration effects compared to healthy controls (Figure 3). As seen in the Table 1, group differences were descriptively more pronounced in the short delay condition where schizophrenia patients had more than two times higher temporal recalibration effects compared to healthy controls on average. We also observed a significant main effect of delay condition, F (1, 38) = 24.71, p < 0.001, η²ₚ = 0.394, reflecting generally greater recalibration in the Long Delay condition compared to the Short Delay condition across participants. However, the interaction between group and delay condition was not significant, F (1, 38) = 0.70, p = 0.408, suggesting that the group difference was similar across conditions and the increase in recalibration with longer delays occurred similarly in both groups. Although the group difference appeared larger in the Short Delay condition descriptively, the non-significant interaction indicates that the schizophrenia group showed similarly elevated temporal recalibration across both delay conditions relative to controls.

**Figure 1:**
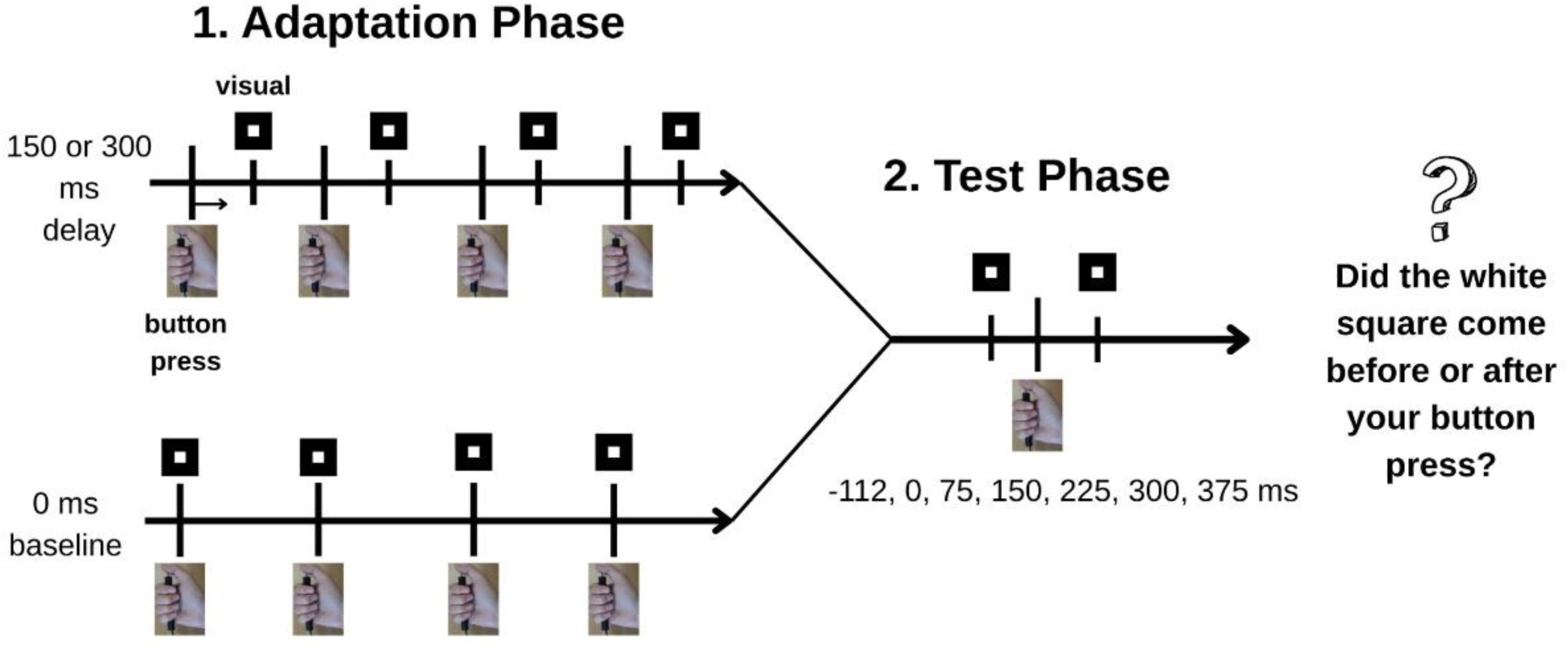
Schematic illustration of the temporal recalibration task. In each trial, participants instructed to perform five consecutive button presses. The first four presses constituted the adaptation phase, during which each press was followed by a visual stimulus (white square) presented at a fixed delay of either 0 ms (baseline condition), 150 ms (short delay condition), or 300 ms (long delay condition). The fifth button press represented the test phase, during which the visual stimulus was presented at one of seven randomly selected stimulus onset asynchronies (SOAs: −112 ms (visual stimulus presented before button press), 0, 75, 150, 225, 300, or 375 ms). Participants made a temporal order judgment, deciding whether the visual stimulus appeared before or after their button press. Conditions were blocked and their order was counterbalanced.

**Figure 2:**
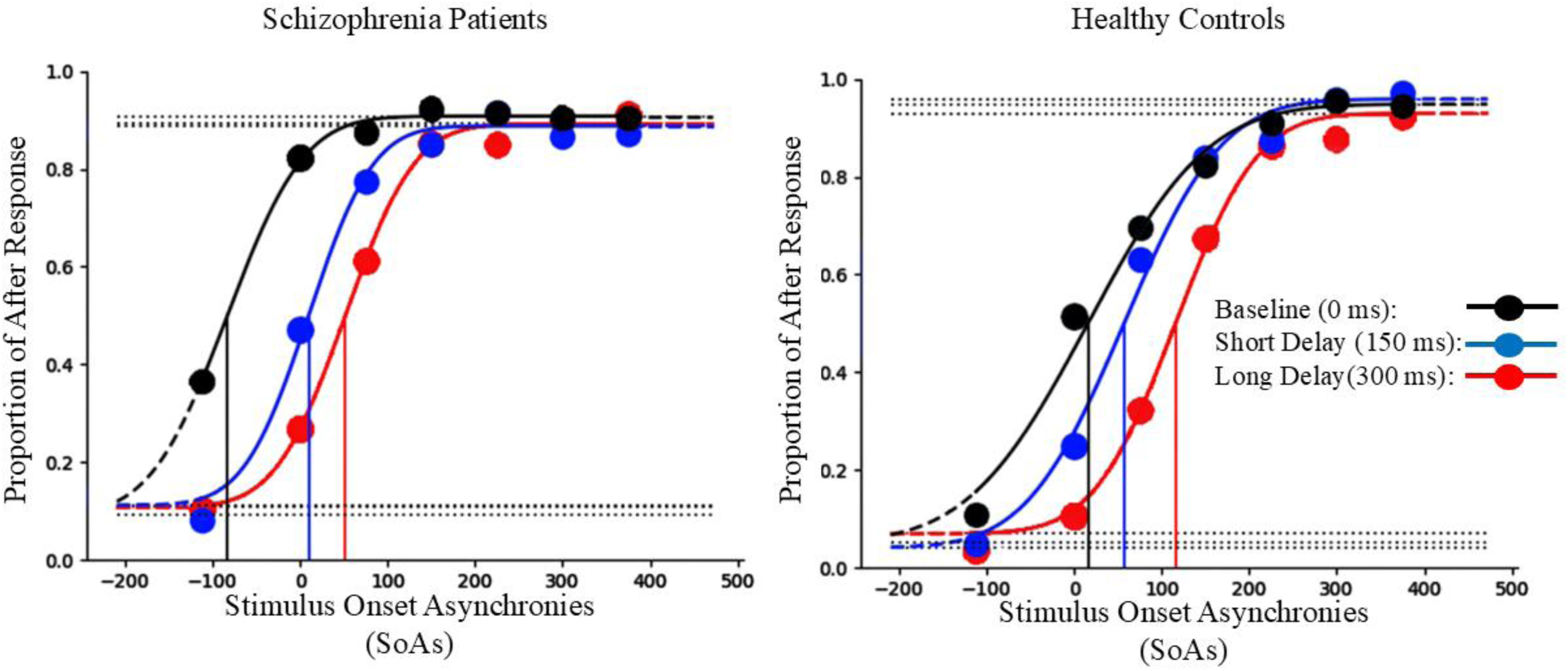
Psychometric functions illustrating temporal order judgments in schizophrenia patients and healthy controls. Each participant’s proportion of “after” responses was plotted as a function of stimulus onset asynchrony (SOA) and fitted using cumulative Gaussian functions. The point of subjective simultaneity (PSS) was defined as the 50% point, where participants were equally likely to judge the visual stimulus as occurring before or after the button press. Functions are presented for the Baseline (0 ms delay; black), Short Delay (150 ms delay; blue), and Long Delay (300 ms delay; red) conditions. Vertical lines indicate the calculated PSS values for each condition, highlighting shifts in perceived simultaneity due to temporal recalibration effects. **Note:** These psychometric functions were derived by aggregating all individual responses within each group (schizophrenia or healthy controls). Specifically, the total number of “after” responses and trials for each SOA were summed across participants within each group, and the resulting group-level data were then fitted. Thus, the PSS values illustrated here reflect group-level fitting and may slightly differ from the mean PSS values presented in Table 1, which were calculated by averaging individual PSS values derived from separately fitted psychometric functions for each participant.

**Figure 3.**
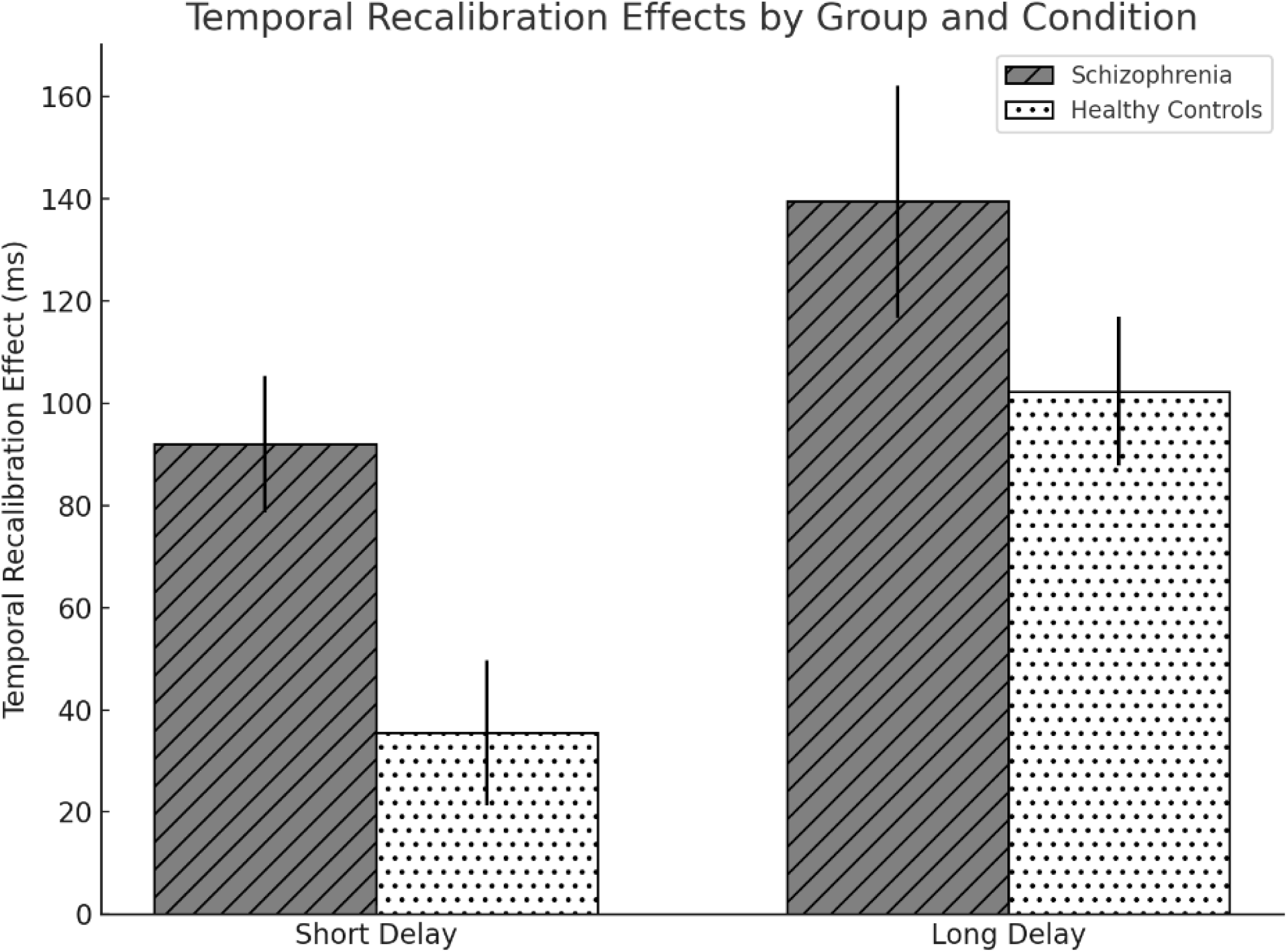
Temporal Recalibration Effects (TRE) Across Groups and Delay Conditions. Bar graph displaying the mean TRE values (± SEM) for schizophrenia patients and healthy controls in the Short Delay and Long Delay conditions. TRE was calculated by subtracting baseline PSS values from the PSS values observed in each delay condition. Main effects of group (p = 0.028) and condition (p < 0.001) were significant. Group x Condition interaction was not significant (p = 0.408).

**Table 1:** Mean (SD) values of point of subjective simultaneity (PSS), just noticeable difference (JND), and temporal recalibration effect (TRE) across conditions for schizophrenia patients and healthy controls.

|  | PSS |  |  | JND |  |  | TRE |  |
| --- | --- | --- | --- | --- | --- | --- | --- | --- |
|  | Baseline | Short | Long | Baseline | Short | Long | Short | Long |
|  |  | Delay | Delay |  | Delay | Delay | Delay | Delay |
| **Schizophrenia** | -67.77 | 24.17 | 71.72 | 88.1 | 113.1 | 111 | 91.94 | 139.48 |
| **Patients** | (83.2) | (64.0) | (102.9) | (76.4) | (95.7) | (63.8) | (59.7) | (101.7) |
| **Healthy** | 22.47 | 58.01 | 124.80 | 93.5 | 94.5 | 94.8 | 35.5 | 102.33 |
| **Controls** | (94.1) | (90.2) | (94.8) | (63.5) | (57.8) | (64.9) | (63.2) | (65.0) |

|  | PSS |  |  | JND |  |  | TRE |  |
| --- | --- | --- | --- | --- | --- | --- | --- | --- |
|  | Baseline | Short | Long | Baseline | Short | Long | Short | Long |
|  |  | Delay | Delay |  | Delay | Delay | Delay | Delay |
| Schizophrenia | -67.77 | 24.17 | 71.72 | 88.1 | 113.1 | 111 | 91.94 | 139.48 |
| Patients | (83.2) | (64.0) | (102.9) | (76.4) | (95.7) | (63.8) | (59.7) | (101.7) |
| Healthy | 22.47 | 58.01 | 124.80 | 93.5 | 94.5 | 94.8 | 35.5 | 102.33 |
| Controls | (94.1) | (90.2) | (94.8) | (63.5) | (57.8) | (64.9) | (63.2) | (65.0) |

As a control analysis, to investigate whether temporal recalibration effect was observed in both group and condition, we conducted a mixed model ANOVA on participants’ PSS thresholds with condition (baseline, short and long delay condition) as within subject and group (schizophrenia patients and healthy controls) as between subject variables. A significant shift of PSS from baseline condition to delay conditions in the direction of the delay would indicate a significant temporal recalibration effect. There was a significant main effect of condition [*F* (2,37) = 40.83, *p* < 0.001, *η²ₚ* = 0.688] and group [*F* (1,38) = 5.69, *p* = 0.022, *η²ₚ* = 0.130]. There was also a significant condition x group interaction effect [*F* (2,37) = 4.13, *p* = 0.024, *η²ₚ* = 0.183]. Pairwise comparisons showed that schizophrenia patients’ PSS threshold significantly shifted from baseline to short delay and to long delay condition (Table 1) (p < 0.001). Similarly, healthy controls’ PSS threshold significantly shifted from baseline to short delay and to long delay condition (p < 0.05). These results suggested that we observed a significant temporal recalibration effect in both groups and conditions. As seen in our previous analysis above, the magnitude of the temporal recalibration effect was greater in the schizophrenia patients.

As another control analysis, a mixed model ANOVA was conducted on the just noticeable difference values (JND) with condition (baseline, short and long delay condition) as within subject and group (schizophrenia patients and healthy controls) as between subject variables. This analysis showed that there was no significant main effect of condition [*F* (2,37) = 0.82, *p* = 0.447] and group [*F* (1,38) = 0.28, *p* = 0.600]. The condition x group interaction effect was also not significant [*F* (2,37) = 0.67, *p* = 0.516]. These results suggest that task difficulty and temporal precision was similar across groups and conditions. Although reduced temporal precision in schizophrenia has been reported, some studies have yielded differential findings.^37,38^ We did not observe lower temporal precision in schizophrenia in our study. This might be due to higher SOA values we used to make the task easier for both groups. Group differences might be observed for lower (more difficult) SOAs. Overall, results of our control analysis suggest that differences in temporal recalibration across groups reflect differential recruitment of this mechanism rather than a general performance deficit in our task.

### Correlations

#### Symptom measures and temporal recalibration

There was no significant correlation between the temporal recalibration effect measures and either SAPS or SANS scores in the schizophrenia group (all |r|s < .25, ps > .25, N = 20), suggesting that temporal recalibration is not directly associated with symptom severity in this clinically stable sample. This null finding may reflect restricted variance in symptom severity, as participants were outpatients with relatively low levels of symptoms, especially with low levels of positive symptoms. Alternatively, temporal recalibration may reflect a more trait-like, compensatory process that operates independently of current symptom levels. Future research should examine this relationship in larger samples with greater variability in clinical status and symptom severity.

#### Baseline PSS and temporal recalibration

Although temporal recalibration is typically defined as the shift in point of subjective simultaneity (PSS) from baseline to delay conditions, we observed baseline PSS differences between groups: schizophrenia patients exhibited more negative PSS values, indicating that the visual stimulus had to be presented earlier to be perceived as simultaneous with the action. This finding aligns with prior research suggesting slowed and impaired visual processing in schizophrenia.^17–21^ To further explore the relationship between baseline PSS and temporal recalibration (i.e., the shift in PSS following exposure to a delay), we conducted an exploratory correlation analysis. In the schizophrenia group, baseline PSS was significantly negatively correlated with temporal recalibration in the short delay condition (r = –0.64, p = .002, N = 20), and showed a weaker, non-significant correlation in the long delay condition (r = –0.39, p = .085, N = 20). Healthy controls showed a similar, though non-significant, pattern: baseline PSS correlated negatively with temporal recalibration in both the short delay (r = – 0.39, p = .081) and long delay (r = –0.34, p = .143) conditions. These findings suggest that greater temporal recalibration in schizophrenia may be partly driven by more negative baseline PSS values, especially under shorter delays. This supports the idea that increased temporal recalibration may serve a compensatory function when baseline sensorimotor synchrony is disrupted. However, the exploratory and correlational nature of these analyses, coupled with the modest sample size, warrants cautious interpretation.

## Discussion

Schizophrenia involves impaired communication and disrupted temporal coordination across widespread brain networks.^1,4,14,15^ Despite these disturbances, patients often maintain coherent perceptual experiences that support adaptive behaviour, particularly during periods of remission. We propose a dual-role hypothesis of temporal recalibration, a process that compensates for temporal discrepancies in sensorimotor integration. Temporal recalibration may support perceptual coherence during stable phases by mitigating neural incoherence, while also potentially contributing to symptom emergence during periods of temporal instability. This hypothesis complements neural network dysconnectivity theories by providing a specific temporal mechanism that could account for the shifts between functional and fragmented perception in schizophrenia. Specifically, we focused on its compensatory role and tested whether clinically stable patients exhibit greater sensorimotor temporal recalibration than healthy controls.

Our results provide empirical evidence that schizophrenia patients demonstrate greater degrees of sensorimotor temporal recalibration in the visuomotor domain. This finding suggests an increased reliance on temporal realignment process to persistent sensorimotor temporal asynchronies. This emphasizes the possible compensatory role of sensorimotor temporal recalibration in counteracting neural network dysconnectivity and facilitating functional perception and effective interaction with the environment in clinically stable patients. Sensorimotor temporal asynchronies requiring increased levels of reliance on compensatory temporal adjustment process might stem from known abnormalities in the timing and integration of wide-spread neural activity. These include impaired efference copy,^14–16^ slowed or less efficient visual processing,^17–21^ reduced connectivity in fronto-occipital network^11^ and cerebellum related dysfunction.^39,40^ These abnormalities may lead to asynchronies across sensorimotor systems that needs to be temporally coordinated by overuse of temporal recalibration. Our finding suggests that schizophrenia patients not only have access to the temporal recalibration mechanism but also may recruit it more than healthy individuals to mitigate disconnection-related delays and preserve temporal coherence. Therefore, temporal recalibration may serve as a temporal correction strategy in schizophrenia during remission.

Our findings can also be interpreted within the predictive processing and Bayesian inference frameworks of schizophrenia. According to this view, increased precision weighting of sensory evidence leads to exaggerated updating of relatively low-precision internal predictions; a biased precision-weighting process thought to be central to perceptual and cognitive disturbances in schizophrenia.^1,41–44^ Temporal recalibration can be conceptualised as a process by which the brain dynamically adjusts its timing expectations in response to repeated exposure to temporal asynchronies.^6,45^ Bayesian views of temporal recalibration suggest that this dynamic process involves updating internal priors to minimize prediction errors between expected and actual temporal relationships.^48,49^ The robust temporal recalibration observed in schizophrenia may reflect the above-described biased precision-weighting. That is, unreliable internal timing predictions are overridden by high-precision incoming evidence, resulting in exaggerated updating of temporal expectations. This is reflected in greater shifts in the point of subjective simultaneity (PSS) based on prior experience and consequently in greater levels of temporal recalibration.

Although increased sensorimotor temporal recalibration may serve an adaptive role, it may also have disruptive consequences. Temporal recalibration studies in healthy individuals show that reducing previously adapted temporal asynchronies can reverse perceived order, making outcomes seem to precede their initiating actions.^6,22,23^ This temporal reversal can disrupt the sense of agency,^24^ the experience of control over our actions and their consequences. As suggested by our findings, dysconnectivity related asynchronies might lead schizophrenia patients to over-rely on recalibration processes to maintain temporal coherence. However, increased levels of temporal recalibration can also increase the risk of experiencing temporal order reversals, especially when transient changes in the adapted asynchronies occur. During acute phases, neural responsivity and excitability can be increased transiently by heightened arousal and neurochemical changes, particularly dopaminergic dysregulation^25–28^ and potentially reduce previously adapted inter-regional temporal relationships. Consequently, processing speeds may temporarily increase and reduce the adapted temporal asynchronies and reverse perceived temporal order. This process may mimic experimental findings in healthy individuals where reducing adapted temporal asynchronies cause outcomes to be perceived as before the actions, thereby disrupting causality and the sense of agency. Consequently, this instability may contribute to the emergence of positive symptoms such as diminished agency, thought insertion, and hallucinations by altering the perceived timing of internally generated experiences. Moreover, temporal recalibration is known to transfer across sensory modalities,^22,23^ and such cross-modal transfer from visual to auditory domains could distort the temporal structure of internal speech. If internal speech or thoughts are perceived as occurring before their initiating intention, they may be experienced as alien or externally inserted. This suggests that exaggerated temporal recalibration in visual domain may also have disruptive effects on auditory domain. Therefore, we suggest that, especially when transient changes in processing speed occur, overreliance on temporal recalibration mechanism has the potential to contribute to positive symptoms and this disruptive effect may generalise to other modalities.

A recent study in schizophrenia patients found that their motor-auditory temporal recalibration was not significantly different than healthy controls.^48^ This contrasts with our findings in the visual modality, where we observed clear group differences. Differential temporal sensitivity across sensory modalities may help explain this discrepancy. Auditory system is known for its superior temporal resolution and faster information processing^49^ which might require less recalibration to maintain smaller asynchronies or may be less affected by internal delays.^46,50^ On the contrary, visual system is slower and more prone to schizophrenia-related delays^17–21^ which can potentially put greater demand for temporal recalibration. Based on these, heightened motor-visual temporal recalibration may reflect a compensatory response to visual timing impairments, which are not as pronounced in the auditory domain. Methodological differences across studies may also contribute to the discrepancy. Schmitter and Straube^48^ employed simultaneity judgement task where the auditory stimulus coincided with or followed the actions, whereas this study used temporal order judgement task which incorporated stimulus onset asynchronies (SOAs) that were physically occurring before the actions. This potentially allowed us to detect negative PSS in the baseline condition of schizophrenia patients. That is, visual stimulus needed to occur earlier to be perceived as simultaneous with the action and this finding is consistent with the visual processing lags in schizophrenia. Future studies investigating both visual and auditory sensorimotor temporal recalibration is needed to uncover the modality specific effects in schizophrenia.

There are limitations to this study that should be acknowledged. First, a methodological limitation concerns the range of SOAs used in the testing phase. In the baseline condition psychometric functions in the schizophrenia group did not reach the lower asymptote whereas this was not the case in healthy control group. This suggests earliest SOA (−112 ms on average) may not have been sufficiently negative to fully capture the PSS in schizophrenia. Consequently, this limitation likely resulted in a less negative baseline PSS than the true value which in turn implies that our estimation of temporal recalibration may be conservative in schizophrenia since temporal recalibration was calculated as the PSS shift from baseline. Therefore, future studies employing wider SOA ranges, particularly with more negative values, may capture more accurate estimation of temporal recalibration in patients. Second, although our sample size is comparable to other studies in the area,^6,7,20,48^ it is modest. Importantly, however, we observed a statistically significant group difference in temporal recalibration, with a moderate-to-large effect size (ηp² = 0.120). This suggests that the observed effect is both robust and potentially meaningful, despite the limited sample. Nevertheless, further studies with larger and more diverse samples are warranted to replicate and extend these findings, and to explore potential moderators such as medication status, symptom severity, or cognitive functioning. Third, in the current study we specifically focused on the visual modality based on previous findings suggesting slower and delayed visual functioning in schizophrenia. However, whether temporal recalibration profiles would be similar in other sensory domains and multisensory contexts needs to be examined. Finally, although neural connectivity problems have been widely shown in the literature and we propose a compensatory role for temporal recalibration in mitigating these, causality cannot be inferred from the present data. This might be tackled with using both neuroimaging and brain stimulation techniques in a similar procedure in the future studies.

## Conclusion

In conclusion, our hypothesis suggests that sensorimotor temporal recalibration may play a previously underrecognized role in schizophrenia, serving both as a compensatory mechanism during remission and a potential contributor to symptom emergence under conditions of temporal instability. We provide evidence that clinically stable patients exhibit exaggerated visuomotor temporal recalibration indicating that the mechanism remains functionally accessible and may be over-engaged to mitigate neural asynchronies. At the same time, we propose that the same mechanism may become maladaptive. When already adapted asynchronies are disrupted, potentially because of transient changes in information processing speed, the temporal recalibration mechanism may distort the perceived order of actions and outcomes. These distortions can contribute to positive symptoms such as hallucinations and delusions of control, particularly by disrupting sense of agency.

This dual-role hypothesis bridges adaptive perceptual timing, predictive processing and clinical symptomology, and offers a novel perspective on how schizophrenia patients may cope with or be destabilized by neural dysconnectivity. Future studies incorporating neural measures, neuromodulation and symptom specific subgroup comparisons may be essential for testing the boundaries and dynamics of this temporal compensation mechanism. Notably, sensorimotor temporal recalibration has been shown to be modifiable by non-invasive brain stimulation such as transcranial direct current stimulation (tDCS) in healthy individuals,^50,51^ highlighting its potential for safe, accessible and targeted modulation. Taken together, these insights may provide a basis for both a mechanistic framework for the temporal organisation of perception in schizophrenia and open new avenues for translational research.

## Data Availability

All data produced in the present study are available upon reasonable request to the authors

## Acknowledgements

This work was supported by Scientific and Technological Research Council of Turkey (TUBITAK) under the Grant Number 122K920. The authors thank to TUBITAK for their supports.

## Author Contribution

Ali A.: Conceptualization, Data Curation, Formal, Analysis, Funding Acquisition, Investigation, Methodology, Project Administration, Resources, Visualisation, Writing-Original draft and Writing- Review and editing. Ayşen E.D.: Data Curation, Investigation and Writing- Review and editing. Osman İ.: Software, Formal Analysis and Writing- Review and editing. All authors approved the final version and submission.

We thank Özüm Karya Sakman for her help with the data collection.

The data that support the findings of this study are available from the corresponding author, A.A., upon reasonable request.

During the preparation of this work, the authors used ChatGPT to improve the text’s language and clarity. After using this tool authors reviewed and edited the content as needed and took full responsibility for the publication’s content.

## Disclosures

Declaration of interest: None

## Notes

### Competing Interest Statement

The authors have declared no competing interest.

### Author Declarations

All procedures involving human subjects/patients were approved by Clinical Research Ethics Committee of the Faculty of Medicine at Manisa Celal Bayar University (Approval No: 429). All participants provided written informed consent prior to participation.

## References

1. Friston K, Brown HR, Siemerkus J, Stephan KE. The dysconnection hypothesis. Schizophr Res. 2016;176(2-3):83–94.

2. Phillips WA, Silverstein SM. Convergence of biological and psychological perspectives on cognitive coordination in schizophrenia. Behav Brain Sci. 2003;26(1):65–82.

3. Ford JM, Mathalon DH. Neural synchrony in schizophrenia. Schizophr Bull. 2008;34(5):904–906.

4. Uhlhaas PJ, Singer W. Abnormal neural oscillations and synchrony in schizophrenia. Nat Rev Neurosci. 2010;11(2):100–113.

5. Fujisaki W, Shimojo S, Kashino M, Nishida SY. Recalibration of audiovisual simultaneity. Nat Neurosci. 2004;7(7):773–778.

6. Stetson C, Cui X, Montague PR, Eagleman DM. Motor-sensory recalibration leads to an illusory reversal of action and sensation. Neuron. 2006;51(5):651–659.

7. Vroomen J, Keetels M, De Gelder B, Bertelson P. Recalibration of temporal order perception by exposure to audio-visual asynchrony. Cogn Brain Res. 2004;22(1):32–35.

8. Andreou C, Faber PL, Leicht G, et al. Resting-state connectivity in the prodromal phase of schizophrenia: insights from EEG microstates. Schizophr Res. 2014;152(2-3):513–520.

9. Assaf R, Ouellet J, Bourque J, et al. Resting-state alterations in emotion salience and default-mode network connectivity in atypical trajectories of psychotic-like experiences. Dev Psychopathol. 2024;1:1.

10. Del Fabro L, Schmidt A, Fortea L, et al. Functional brain network dysfunctions in subjects at high-risk for psychosis: a meta-analysis of resting-state functional connectivity. Neurosci Biobehav Rev. 2021;128:90–101.

11. Khadka S, Meda SA, Stevens MC, et al. Is aberrant functional connectivity a psychosis endophenotype? A resting state functional magnetic resonance imaging study. Biol Psychiatry. 2013;74(6):458–466.

12. Pelletier-Baldelli A, Andrews-Hanna JR, Mittal VA. Resting state connectivity dynamics in individuals at risk for psychosis. J Abnorm Psychol. 2018;127(3):314.

13. Wang C, Ji F, Hong Z, et al. Disrupted salience network functional connectivity and white-matter microstructure in persons at risk for psychosis: findings from the LYRIKS study. Psychol Med. 2016;46(13):2771–2783.

14. Whitford TJ, Ford JM, Mathalon DH, Kubicki M, Shenton ME. Schizophrenia, myelination, and delayed corollary discharges: a hypothesis. Schizophr Bull. 2012;38(3):486–494.

15. Parlikar TA, Cahill CM, Whitford TJ. Altered myelination as a contributor to schizophrenia: Evidence from diffusion tensor imaging studies. Front Psychiatry. 2019;10:173.

16. Pynn LK, DeSouza JF. The function of efference copy signals: Implications for symptoms of schizophrenia. Vision Res. 2013;76:124–133.

17. Adámek P, Langová V, Horáček J. Early-stage visual perception impairment in schizophrenia, bottom-up and back again. Schizophrenia. 2022;8(1):27.

18. Francisco AA, Foxe JJ, Horsthuis DJ, Molholm S. Early visual processing and adaptation as markers of disease, not vulnerability: EEG evidence from 22q11.2 deletion syndrome, a population at high risk for schizophrenia. Schizophrenia. 2022;8(1):28.

19. Jiang CG, Wang J, Liu XH, Xue YL, Zhou ZH. The Neural Correlates of Effortful Cognitive Processing Deficits in Schizophrenia: An ERP Study. Front Hum Neurosci. 2021;15:664008.

20. Johnson SC, Lowery N, Kohler C, Turetsky BI. Global–local visual processing in schizophrenia: evidence for an early visual processing deficit. Biol Psychiatry. 2005;58(12):937–946.

21. Silverstein S, Keane BP, Blake R, et al. Vision in schizophrenia: why it matters. Front Psychol. 2015;6:41.

22. Heron J, Hanson JV, Whitaker D. Effect before cause: supramodal recalibration of sensorimotor timing. PLoS One. 2009;4(11):e7681.

23. Sugano Y, Keetels M, Vroomen J. Adaptation to motor-visual and motor-auditory temporal lags transfer across modalities. Exp Brain Res. 2010;201:393–399.

24. Timm J, Schönwiesner M, SanMiguel I, Schröger E. Sensation of agency and perception of temporal order. Conscious Cogn. 2014;23:42–52.

25. Corlett PR, Fraser KM. 20 years of aberrant salience in psychosis: What have we learned? Am J Psychiatry. 2025;appiajp.

26. Grace AA. Dopamine system dysregulation by the hippocampus: implications for the pathophysiology and treatment of schizophrenia. Neuropharmacology. 2012;62(3):1342–1348.

27. Lodge DJ, Grace AA. Hippocampal dysregulation of dopamine system function and the pathophysiology of schizophrenia. Trends Pharmacol Sci. 2011;32(9):507–513.

28. McCutcheon RA, Krystal JH, Howes OD. Dopamine and glutamate in schizophrenia: biology, symptoms and treatment. World Psychiatry. 2020;19(1):15–33.

29. First MB. Structured Clinical Interview for DSM-IV Axis I Disorders (SCID-I), Clinician Version (Administration Booklet). Washington, DC: American Psychiatric Publishing, Inc.; 1997.

30. Andreasen NC. The Scale for the Assessment of Positive Symptoms (SAPS). Iowa City, IA: University of Iowa; 1984.

31. Erkoç Ş, Arkonaç O, Ataklı C, Özmen E. Pozitif semptomları değerlendirme ölçeğinin güvenilirliği ve geçerliliği. Düşünen Adam. 1991;4(2):20–24.

32. Andreasen NC. The Scale for the Assessment of Negative Symptoms (SANS). Iowa City, IA: University of Iowa; 1984.

33. Erkoç Ş, Arkonaç O, Ataklı C, Özmen E. Negatif semptomları değerlendirme ölçeğinin güvenilirliği ve geçerliliği. Düşünen Adam. 1991;4(2):14–15.

34. Mathôt S, Schreij D, Theeuwes J. OpenSesame: An open-source, graphical experiment builder for the social sciences. Behav Res Methods. 2012;44(2):314–324.

35. Schubert TW, D’Ausilio A, Canto R. Using Arduino microcontroller boards to measure response latencies. Behav Res Methods. 2013;45:1332–1346.

36. Schütt HH, Harmeling S, Macke JH, Wichmann FA. Painfree and accurate Bayesian estimation of psychometric functions for (potentially) overdispersed data. Vis Res. 2016;122:105–123.

37. Martin B, Giersch A, Huron C, van Wassenhove V. Temporal event structure and timing in schizophrenia: preserved binding in a longer “now”. Neuropsychologia. 2013;51(2):358–371.

38. Thoenes S, Oberfeld D. Meta-analysis of time perception and temporal processing in schizophrenia: Differential effects on precision and accuracy. Clin Psychol Rev. 2017;54:44–64.

39. Andreasen NC, Pierson R. The role of the cerebellum in schizophrenia. Biol Psychiatry. 2008;64(2):81–88.

40. Kim SE, Jung S, Sung G, Bang M, Lee SH. Impaired cerebro-cerebellar white matter connectivity and its associations with cognitive function in patients with schizophrenia. NPJ Schizophr. 2021;7(1):38.

41. Adams RA, Stephan KE, Brown HR, Frith CD, Friston KJ. The computational anatomy of psychosis. Front Psychiatry. 2013;4:47.

42. Clark A. Whatever next? Predictive brains, situated agents, and the future of cognitive science. Behav Brain Sci. 2013;36(3):181–204.

43. Fletcher PC, Frith CD. Perceiving is believing: a Bayesian approach to explaining the positive symptoms of schizophrenia. Nat Rev Neurosci. 2009;10(1):48–58.

44. Sterzer P, Adams RA, Fletcher P, et al. The predictive coding account of psychosis. Biol Psychiatry. 2018;84(9):634–643.

45. Parsons BD, Novich SD, Eagleman DM. Motor-sensory recalibration modulates perceived simultaneity of cross-modal events at different distances. Front Psychol. 2013;4:46.

46. Di Luca M, Machulla TK, Ernst MO. Recalibration of multisensory simultaneity: cross-modal transfer coincides with a change in perceptual latency. J Vis. 2009;9(12):7.

47. Sadibolova R, Terhune DB. The temporal context in bayesian models of interval timing: Recent advances and future directions. Behav Neurosci. 2022;136(5):364.

48. Schmitter CV, Straube B. Facilitation of sensorimotor temporal recalibration mechanisms by cerebellar tDCS in patients with schizophrenia spectrum disorders and healthy individuals. Sci Rep. 2024;14(1):2627.

49. Kanai R, Lloyd H, Bueti D, Walsh V. Modality-independent role of the primary auditory cortex in time estimation. Exp Brain Res. 2011;209:465–471.

50. Aytemür A, Almeida N, Lee KH. Differential sensory cortical involvement in auditory and visual sensorimotor temporal recalibration: Evidence from transcranial direct current stimulation (tDCS). Neuropsychologia. 2017;96:122–128.

51. Schmitter CV, Straube B. The impact of cerebellar transcranial direct current stimulation (tDCS) on sensorimotor and inter-sensory temporal recalibration. Front Hum Neurosci. 2022;16:998843.

