## Supplementary Table 1 for "Temporal Recalibration in Schizophrenia: A Compensatory Timing Trap?"

| Variable | Schizophrenia (n = 20) | Healthy Controls (n = 20) |
| --- | --- | --- |
| Gender (Male %) | 60% | 60% |
| Age, M (SD) | 38.90 (9.79) | 39.60 (9.29) |
| Education (years), M (SD) | 13.00 (3.93) | 11.85 (3.36) |
| Socioeconomic Status, M (SD) | 4.65 (1.57) | 5.75 (1.41) |
| SAPS Total, M (SD) | 6.20 (6.92) | - |
| SANS Total, M (SD) | 15.45 (14.08) | - |
| Hallucinations, M (SD) | 1.25 (2.51) | - |
| Delusions, M (SD) | 1.65 (2.35) | - |
| Bizarre Behavior, M (SD) | 0.80 (1.20) | - |
| Formal Thought Disorder, M (SD) | 2.50 (2.89) | - |
| Affective Flattening, M (SD) | 3.50 (4.97) | - |
| Alogia, M (SD) | 1.80 (2.78) | - |
| Avolition, M (SD) | 2.40 (2.64) | - |
| Anhedonia-Asociality, M (SD) | 5.20 (4.69) | - |
| Attention, M (SD) | 2.55 (2.35) | - |

Supplementary Table 1 Demographic and Clinical Characteristics of the Schizophrenia and Control Groups
